# Real-World Experience of Deep Brain Stimulation Surgery in a Developing Southeast Asian Country

**DOI:** 10.1101/2023.08.23.23294286

**Authors:** Alfand Marl F. Dy Closas, Ai Huey Tan, Yi Wen Tay, Jia Wei Hor, Tzi Shin Toh, Jia Lun Lim, Choey Yee Lew, Chun Yoong Cham, Carolyn Chue Wai Yim, Kok Yoon Chee, Chong Guan Ng, Lei Cheng Lit, Anis Nadhirah Khairul Anuar, Lara M. Lange, Zih-Hua Fang, Sara Bandres Ciga, Katja Lohmann, Christine Klein, Azlina Ahmad-Annuar, Kalai Arasu Muthusamy, Shen-Yang Lim

## Abstract

**Background:** The availability of deep brain stimulation (DBS), a highly efficacious treatment for several movement disorders, remains low in developing countries, with scarce data available on utilization and outcomes.

**Objectives:** We characterized the DBS cohort and outcomes at a Malaysian quaternary medical centre.

**Methods:** A retrospective chart review was done on DBS-related surgery at the University of Malaya, including clinico-demographic, genetics, and outcomes data focusing on post-operative medication reduction and complications.

**Results:** 149 Parkinson’s disease (PD) patients underwent DBS targeting the subthalamic nucleus. Six had globus pallidus internus DBS (primarily for dystonia). Only 16.1% of cases were government-funded. Of the 133 PD patients operated in the past decade (2013-2022), 25 (18.8%) had disease duration <5 years. At 6-12 months post-DBS, median levodopa-equivalent daily dosage (LEDD) reduction was 440.5 [418.9] mg/day, corresponding to a reduction of ≥50% and ≥30% in 42.2% and 69.8% of patients, respectively. LEDD reductions were larger in the early-onset and short-duration subgroups. Three patients (1.9% of 155) had symptomatic intracranial hemorrhage, resulting in stroke in two. Pathogenic monogenic or *GBA1* variants were detected in 12/61 (19.7%) of patients tested, mostly comprising the “severe” *GBA1* variant p.L483P (14.8%).

**Conclusion:** This is the largest report on DBS from Southeast Asia. The procedures were effective, and complication rates on par with international norms. Our study found a high frequency of *GBA1*-PD; and included a substantial number of patients with short-duration PD, who had good outcomes. It also highlights the inequity of access to device-aided therapy.

## 1. INTRODUCTION

Deep brain stimulation (DBS) is an established treatment for Parkinson’s disease (PD) and other movement disorders, often dramatically improving motor functions in treated patients, with sustained benefits reported for 10 years or more.^1,2^ Since its introduction into clinical practice in the late 1990s, advances in hardware, software, and targeting have further improved treatment efficacy and device functionality.^3^ Patient selection has become more inclusive with expanded disease indications, and a trend towards offering the procedure earlier in the disease course.^2–4^ The EARLYSTIM trial^4^ published a decade ago showed that DBS performed earlier in the course of PD (trial patients were aged ≤60 years, with disease duration ≥4 years and ≤3 years of motor response complications) was effective and safe. The publication of this seminal study, together with the advent of longer-life implantable pulse generators (IPGs; lasting ≥15 years) have likely changed the landscape of DBS application worldwide,^5,6^ but few studies have documented this important shift in real-world clinical practice.^7,8^ Concerns also continue to be put forth in opposition to this approach, including, understandably, the possible misdiagnosis of early-stage atypical parkinsonism for PD.^9,10^

Furthermore, because the availability of DBS in developing countries remains low, due to its high cost, and limitations in government healthcare expenditure, facilities and expertise,^11,12^ there is a paucity of data on DBS utilization and outcomes, e.g., from Southeast Asia with a population of >650 million (total *n*=219 patients across 10 studies, sample sizes ranging from 1 to 56 patients – references in the Supplementary materials Appendix A). The cost of other device-aided therapies (in particular, infusions of dopaminergic agents) is even more prohibitive in this region, making them even less available/accessible to patients.^12,13^ Indeed, the lack of access to these potentially life-changing treatments in low– and middle-income countries – where most people affected by PD reside – has been highlighted by the World Health Organization as an area of health disparity in need of global action.^14^

There has also been increasing interest in exploring genotype-phenotype correlations, including the influence of genetic variants on the disease course of PD, dystonias, and other movement disorders, and their response to DBS.^15–19^ In the era of personalized precision medicine, a greater understanding of the role of genetics can help to refine the selection of treatments, for optimized patient outcomes.^18–21^

In this study, we aimed to characterize the clinico-demographic features of our DBS cohort and their outcomes, at a quaternary medical centre in Malaysia. The results of genetic testing, where available, were reviewed.

## 2. METHODS

### 2.1 Subjects, deep brain stimulation procedure, and data collection

A retrospective chart review was done on consecutive patients undergoing DBS surgeries at the University of Malaya (UM), since its inception in September 2004, until December 2022. We included patients who underwent new/primary DBS implantations, as well as those having IPG replacement or revision surgeries. The DBS protocol at UM including pre-operative workup, surgical procedure, and post-DBS management are detailed in the Supplementary materials Appendix B. The vast majority (>90%) of patients were managed by either one of two movement disorder neurologists (SYL, AHT) and one DBS neurosurgeon (KAM).

Clinico-demographic and outcomes data focusing on post-operative PD medication reduction and complications were collected. Patients were managed pragmatically, rather than in a research setting, and motor improvement post-operatively was usually assessed qualitatively, in a variety of (non-uniform) ways, including reduction in the severity and duration of OFF periods, dyskinesias and/or tremors, and improvements in functional abilities or quality of life. Patients were rarely taken off their PD medications after completion of DBS programming to formally measure OFF-medication, ON-DBS status (this was only done if there was uncertainty about a lack of efficacy from stimulation). Thus, in this study, post-operative reduction in PD medications (calculated as levodopa-equivalent daily dose [LEDD]^22,23^) was used as a surrogate measure for motor improvement after subthalamic nucleus (STN) DBS. LEDDs were calculated at the following time points: (i) pre-DBS (T1); (ii) within 6-12 months after surgery (T2); and (iii) at the most recent hospital visit (more than 12 months after surgery) (T3). In the published literature, LEDD reduction after STN DBS has typically been ≍30-50%,^2,24,25^ with a trend for increase (≈10%) on prolonged follow-up.^2,25^ Rating scales were not systematically administered in the small group of dystonia patients pre– and post-DBS, and the benefit from DBS was rated qualitatively by SYL (small, medium, or large).

We also examined the proportion and outcomes of patients undergoing DBS with short duration or early onset of PD. Given that guidelines commonly recommend that ≥5 years have elapsed from disease onset before performing DBS (henceforth referred to as “standard-duration” PD) in order to avoid inclusion of atypical parkinsonian disorders,^5,9,26^ here we defined “short-duration” PD patients as those undergoing DBS <5 years after diagnosis. Early-onset PD (EOPD) was defined as those diagnosed ≤50 years of age, and late-onset PD (LOPD) were diagnosed after age 50 years.

The validated results of genetic tests were reviewed. These were done on a research basis, and in almost all cases the results only became available after DBS surgery had already been performed. Ethical approval was obtained from the Medical Ethics Committee, UM.

### 2.2 Statistical analyses

Descriptive statistics were used to analyse clinical and demographic parameters. Shapiro-Wilk test was used for normality testing. Wilcoxon signed rank test was used to analyze the pairwise differences in the LEDD at different timepoints and across sub-groups. Mann-Whitney U test was used to compare the quantitative clinico-demographic parameters and LEDD reduction between cohort subgroups. Chi-square test was used to analyze the differences in the qualitative demographic characteristics between subgroups and distributions of % LEDD reduction category between subgroups. Patients who had their DBS system removed or were off stimulation were still included in the analysis if data were available.

## 3. Results

### 3.1 DBS procedures performed

There were 161 patients who underwent DBS-related surgeries (314 leads implanted, 4 leads re-positioned, 175 IPGs implanted [154 newly implanted, 21 replaced after “end of service”]) (Figure 1 and Table 1). Of the 155 patients who underwent the primary bilateral DBS surgery at UM, 147 (94.8%) had synchronous bilateral STN DBS; two (1.3%) had staged bilateral STN DBS (one because of subdural haematoma occurring intra-operatively; and one, a patient with severe left-sided PD features, initially underwent right-sided STN DBS in 2005 using a Medtronic Soletra^®^ IPG); and six (3.9%) had synchronous bilateral globus pallidus internus (GPi) DBS. Of the 149 PD patients whose first DBS surgery (DBS leads and IPG implantation) done at UM, only one, who had severe dyskinesias even with very low-dosage dopamine agonist therapy, underwent DBS targeting the GPi. The remaining five patients who had GPi DBS had dystonia (of whom, one had combined dystonia-parkinsonism). There were no cases of thalamic DBS for tremor disorders.

**Figure 1.**
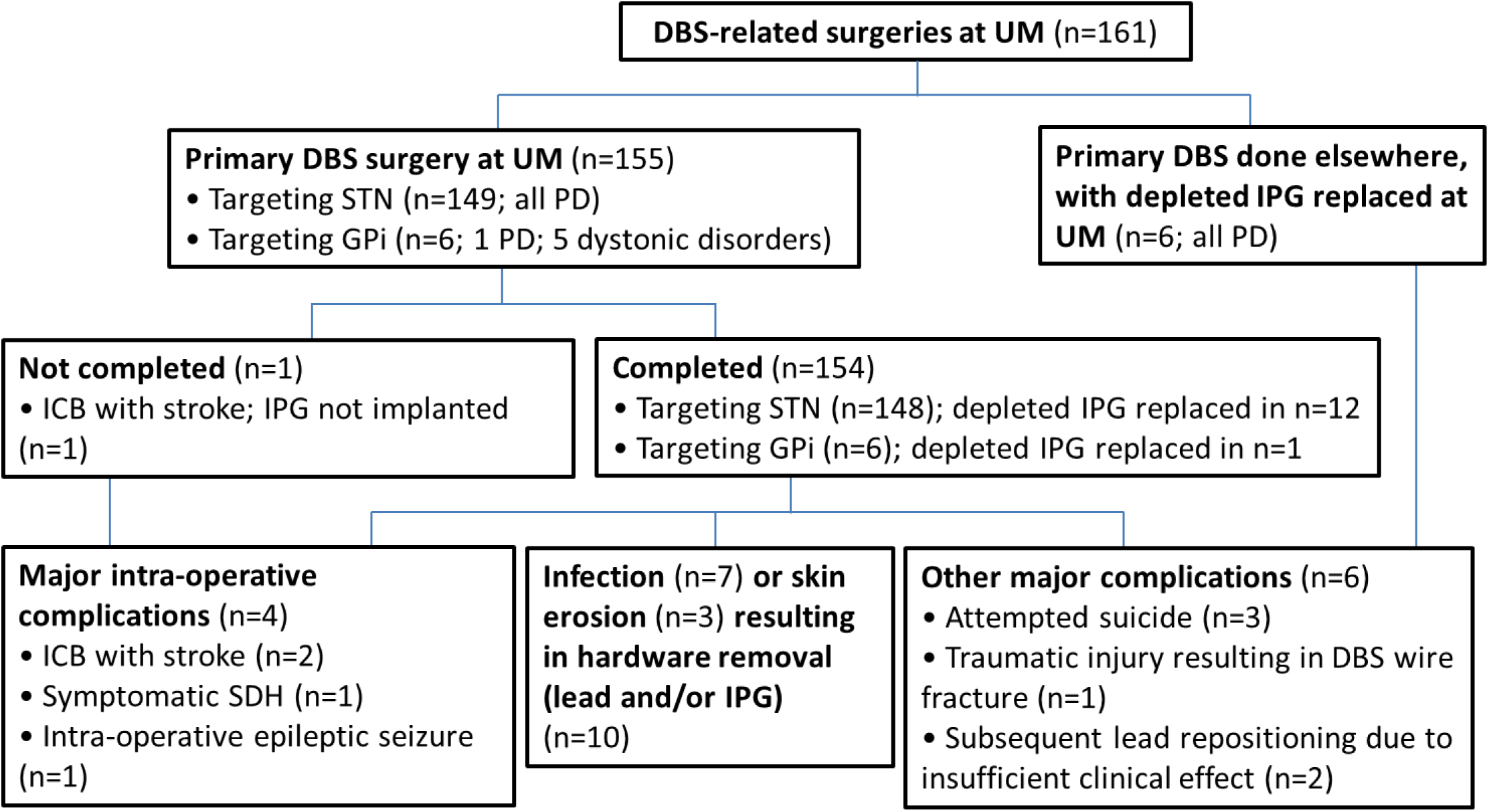
Patient flow diagram. Abbreviations: ICB=Intracerebral bleeding; n=Number of patients; SDH=Subdural haematoma; UM=University of Malaya

**Table 1.** Clinico-demographic data of patients undergoing deep brain stimulation-related surgeries at the University of Malaya.

| PARAMETER | Overall cohort (n=161) | PD subgroup (n=156) | Early-onset PD subgroup (n=85) | Late-onset PD subgroup (n=71) | Duration <5y subgroup (n=25) | Duration ≥5y subgroup (n=131) |
| --- | --- | --- | --- | --- | --- | --- |
| Age at surgery (years) | 58.7 [11.9] | 58.9 [11.8] | 53.2 [8.6] | 64.1 [6.3] | 57.5 [16.3] | 59.6 [11.0] |
| Male (%) | 61.5 | 59.0 | 68.1 | 52.9 | 52.0 | 61.6 |
| Ethnicity (%) |  |  |  |  |  |  |
| Chinese | 73.9 | 74.4 | 73.6 | 74.3 | 64.0 | 76.8 |
| Indian | 13.7 | 13.5 | 13.2 | 14.3 | 12.0 | 13.6 |
| Malay | 6.2 | 5.8 | 7.7 | 4.3 | 16.0 | 4.0 |
| Others | 6.2 | 6.4 | 5.5 | 7.1 | 8.0 | 5.6 |
| Disease duration (years) | 8.8 [5.9] | 8.8 [5.7] | 9.7 [6.6] | 7.0 [4.8] | 3.4 [1.4] | 9.7 [5.4] |
| Very short duration (<4y) |  | 16 (10.3%) |  |  |  |  |
| Short duration (4 to <5y) |  | 9 (5.8%) |  |  |  |  |
| Age at diagnosis | 49.0 [13.0] | 49.0 [13.0] | 43.0 [9.0] | 56.0 [6.0] | 54.0 [17.0] | 48.0 [13.0] |
| Young-onset (≤40 years) |  | 28 (17.9%) |  |  |  |  |
| Early-onset (≤50 years) |  | 85 (54.5%) |  |  |  |  |
| Late-onset (>50years) |  | 71 (45.5%) |  |  |  |  |
| Implantation technique (n=155) |  |  |  |  |  |  |
| Bilateral synchronous | 153 | 148 |  |  |  |  |
| Bilateral staged | 2 | 2 |  |  |  |  |
| DBS Target (n=155) |  |  |  |  |  |  |
| STN | 149 | 149 |  |  |  |  |
| GPi | 6 | 1 |  |  |  |  |
| Number of DBS leads (n=314) |  |  |  |  |  |  |
| Newly implanted | 310 |  |  |  |  |  |
| Repositioned | 4 |  |  |  |  |  |
| Explanted and replaced | 4 |  |  |  |  |  |
| Explanted, not replaced<sup>a</sup> | 15 |  |  |  |  |  |
| Number of IPGs (n=175) |  |  |  |  |  |  |
| Newly implanted | 154 |  |  |  |  |  |
| Replaced<sup>a</sup> | 21 |  |  |  |  |  |
| Repositioned | 3 |  |  |  |  |  |
| Explanted, not replaced | 5 |  |  |  |  |  |

Dystonia patients who comprised only a small subgroup are described in the manuscript text and not tabulated here. Values are presented as mean ± standard deviation where data were normally distributed, otherwise, as median [interquartile range (IQR)].

The number of primary DBS surgeries performed have increased over time, except for the period 2020-2021, due to the COVID-19 pandemic (Figure 2). On average, 19 primary DBS surgeries were performed annually from 2016-2022.

**Figure 2.**
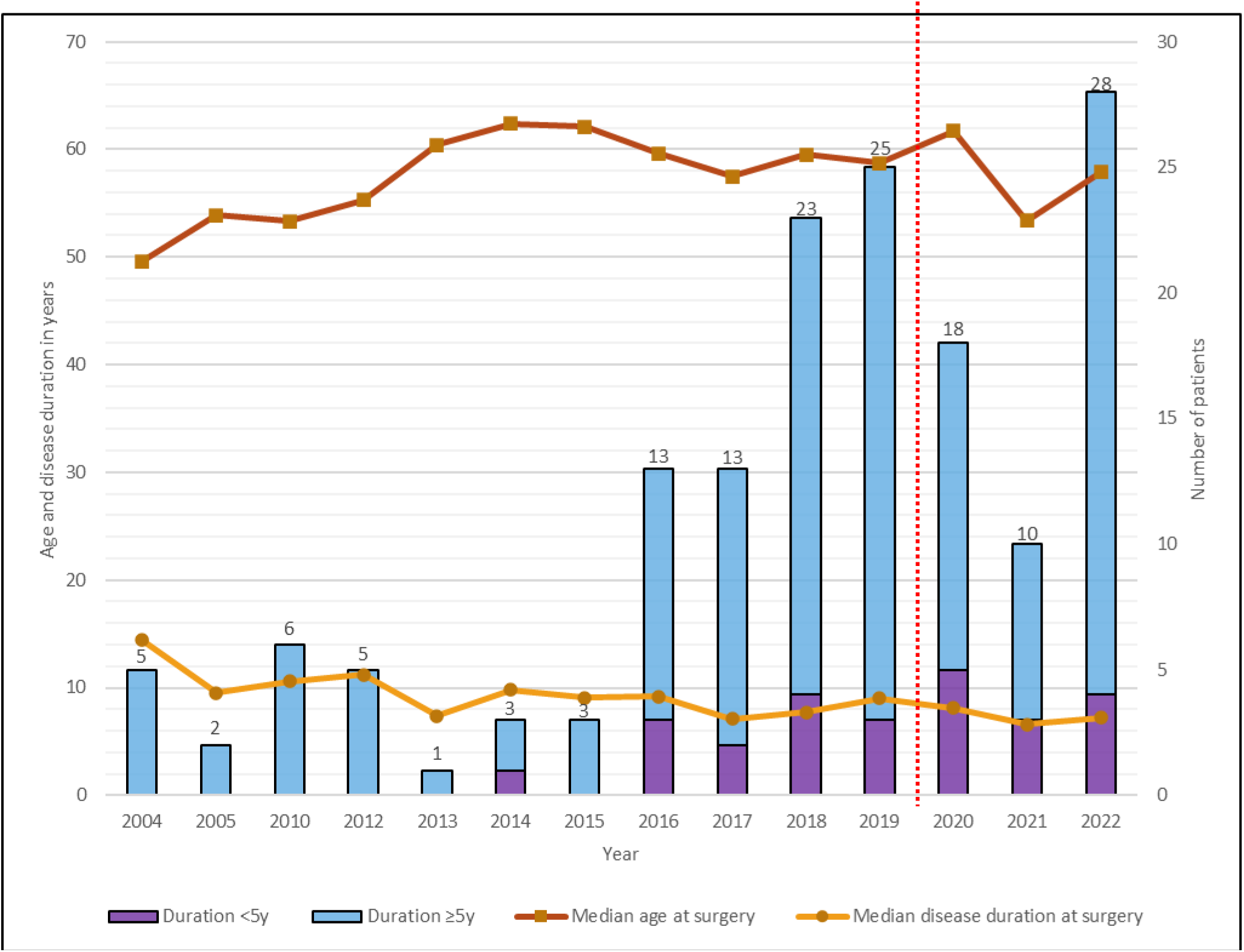
Median disease duration and age at first/primary deep brain stimulation (DBS) surgery for Parkinson’s disease (PD), and the number of patients with short-duration and standard-duration PD from 2004-2022. Short-duration PD patients were defined as those undergoing DBS <5 years after PD diagnosis; while standard-duration PD was defined as ≥5 years. The red dotted line represents the start of COVID-related restrictions.

The vast majority of the DBS surgeries were self-paid or reimbursed by private health insurance; only 26 cases (16.1%, patients who were government-employed) received government funding.

The median interval between surgery and first DBS programming was 5.4 weeks [IQR: 1.9; range 2.7-20.4 weeks]. The duration of post-operative follow-up was 2.9 [3.5] years (0-17.4 years). The majority of the cohort were still under active follow-up; 24 (14.9%) were lost to follow-up and another 16 (9.9%) were deceased at T3 (age and disease duration at death 70 [8.6] and 17.5 [9.7] years, respectively). One patient died within the first post-operative year (41 weeks post-DBS), from cardiac arrhythmia related to hypokalemia.

### 3.2 Surgery and hardware-related complications

Among the 155 patients who had their primary DBS surgery at UM, two (1.3%) developed large intracerebral (frontal lobe) bleeds intra-operatively causing stroke, with disabling and persisting neurological deficits (Figure 1 and Supplementary Table 1). Another patient with intra-operative subdural hematoma causing severe headache underwent immediate hematoma evacuation without neurological deficits. One with occipital lobe infarction pre-operatively had intra-operative seizure (0.6%).

In the overall cohort (n=161), seven (4.3%) and three patients (1.9%) developed surgical site infection (SSI) and skin erosion, respectively (Supplementary Table 1). Despite antibiotic treatment and surgical debridement, explantation of the DBS leads and/or IPG was eventually needed in six patients. Underlying dermatological conditions led to SSI in two of them (one with drug-induced Stevens-Johnson syndrome, another with bullous pemphigoid and diabetes mellitus).

Two patients had bilateral lead repositioning for suboptimal stimulation effect. Additionally, three patients underwent re-implantation of DBS leads: two for infection (bilateral STN leads in one, and unilateral STN lead in another who initially had DBS in another institution), and one for lead fracture after a major fall (six years after the initial DBS surgery, in a patient with short-duration PD).

### 3.3 PD patients

There were 156 patients with PD, the majority male and Chinese (61.5% and 73.9%, respectively; Table 1), closely comparable to the overall PD demographic at UM.^27^ The median ages at diagnosis and at first DBS surgery were 49.0 [13.0] (range: 25.0-68.0 years) and 58.9 [11.8] (35.8-77.3) years, respectively, with disease duration from diagnosis of 8.8 [5.7] (range: 2.3-24.3) years. EOPD cases (n=85) comprised 54.5%, of whom 28 (17.9% of the overall PD cohort) were young-onset (age at diagnosis ≤40 years). LOPD cases comprised the remaining 45.5% (n=71) of the cohort.

Over time, there was a trend for reducing PD duration in patients undergoing their first DBS surgery (Figure 2), with increasing numbers, since around 2014, of patients with short-duration PD. Disease duration from diagnosis was <5 years in 25 patients (age at surgery: 59.6 [11.0] years; disease duration: 3.4 [1.4] years), comprising 18.8% of the 133 PD patients operated in the past decade (2013-2022). As was the case for the overall PD group, troublesome motor response complications were the usual primary indication for DBS (Supplementary Table 2), with only a few patients having troublesome medication-unresponsive tremor. None had a change in diagnosis to an atypical parkinsonian disorder over follow-up (2.3 [2.6], range: 0-5.6 years).

In the overall PD cohort, the median reduction in LEDD at T2 (vs. T1) was 440.5 [418.9] mg/day (P<0.001; n=116), with 42.2% (n=49) and 69.8% (n=81) of patients having ≥50% and ≥30% reductions, respectively (Figure 3, A-C). The median LEDD reduction at T3 (vs. T1) was 297.6 [494.3] mg/day (P<0.001; n=110), with 30.0% (n=33) and 58.2% (n=64) having ≥50% and ≥30% reductions (Figure 3, A, B and D). Median LEDD was significantly higher at T3 vs. T2 (P=0.007).

**Figure 3.**
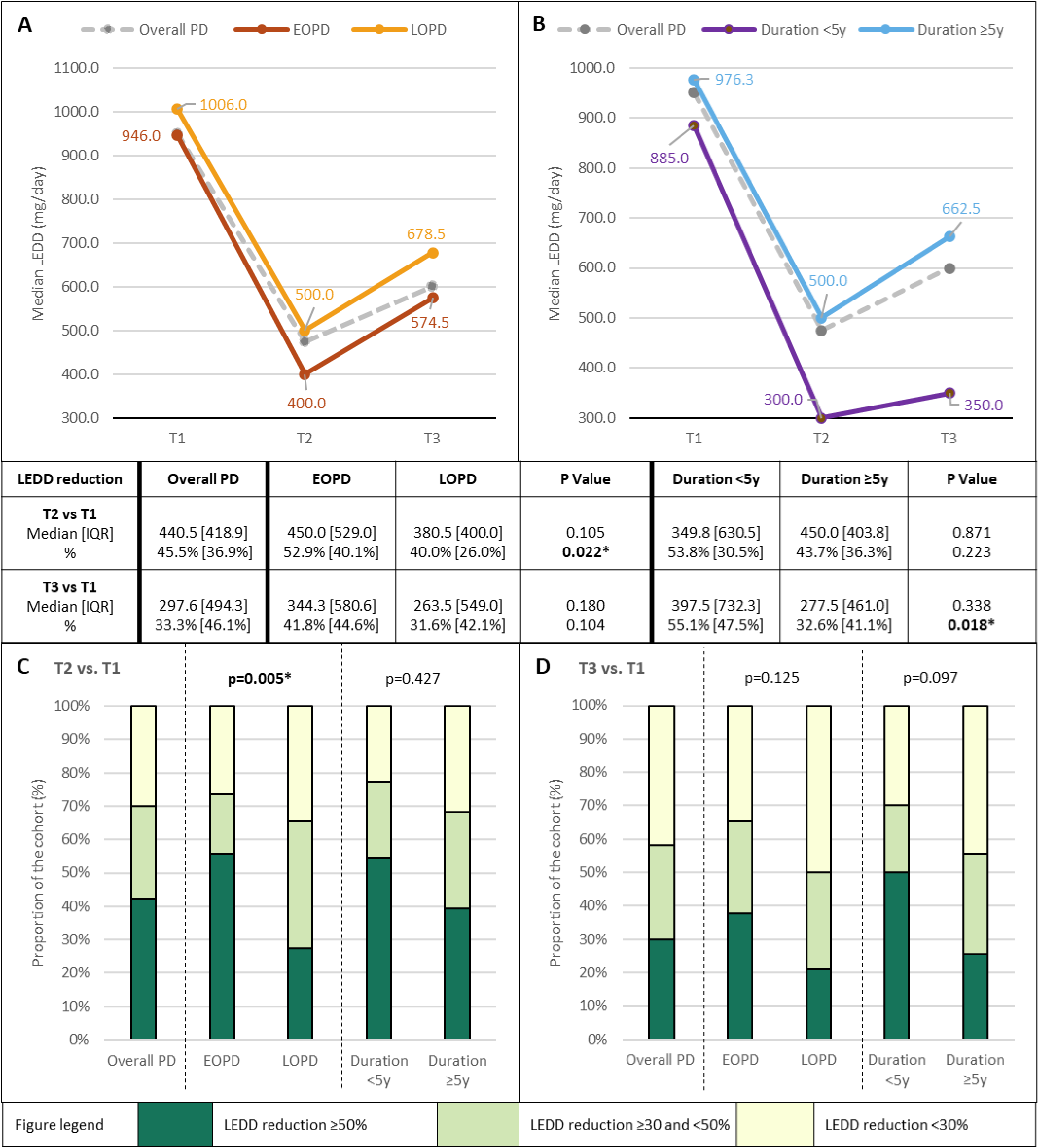
Changes in levodopa-equivalent daily dosage (LEDD) after deep brain stimulation (DBS) surgery. Median changes in LEDD at T2 and T3 (both vs. T1) are depicted in A and B, and the proportional distributions of LEDD reductions (≥50%; ≥30 and <50%; <30%) at T2 in C and at T3 in D. Comparisons between early (≤50y)-vs. late (>50y)-onset PD subgroups are depicted in A, C and D, and between short (<5y)-vs. standard (≥5y)-duration PD in B, C and D. T1=Pre-DBS; T2=Within 6-12 months post-DBS; T3=At last hospital visit. *Denotes significant difference.

In the subgroup analyses of EOPD and short-duration patients: (i) the T2-vs.-T1 median % LEDD reduction was significantly greater in the EOPD vs. LOPD subgroup (52.9% [40.1%] vs. 40.0% [26.0%], P=0.022) with a significantly higher proportion of EOPD patients with ≥50% LEDD reduction (55.7% vs. 27.3%; P=0.005) (Figure 3, A and C); and (ii) the T3-vs.-T1 median % LEDD reduction was significantly greater in the short-vs. standard-duration PD subgroup (55.1% [47.5%] vs. 32.6% [41.1%], P=0.018), with a trend for a higher proportion of short-duration patients with ≥50% LEDD reduction (50.0% vs 25.6%, P=0.097) (Figure 3, B and D). Follow-up duration was not significantly different between the short-vs. standard-duration PD groups (2.5 [2.4] years vs. 2.9 [3.5] years, P=0.567).

Regarding neuropsychiatric problems post-DBS, suicide was attempted in three patients (1.9%) while four patients displayed clinically-overt impulsive-compulsive behaviors (ICBs). Four patients (2.6%) had new-onset psychosis within the first post-operative year, one of whom required urgent psychiatry referral. Another patient with pre-existing psychosis had worsening psychosis post-DBS that also required urgent psychiatry care. Twelve patients (7.5%) went on to develop dementia after a median of 4.6 years after DBS. Age at the last visit (67.5 [38.1] vs. 62.5 [52.8] years, P=0.036) and disease duration (15.3 [27.8] vs. 11.3 [31.5] years, P=0.031) were significantly higher/longer in those with dementia vs. those without. Only one patient of 12 who developed dementia came from the short-duration PD subgroup.

Results of testing for monogenic and *GBA1*-related PD (which prioritized familial and/or EOPD cases) were available for 61 patients: next generation sequencing-based PD gene panel (n=21),^15^ multiplex ligation-dependent probe amplification (n=13),^28^ and/or whole genome sequencing (n=44, under the Global Parkinson’s Genetics Project [GP2]).^17^ Twelve (19.7%) were found to have variants in the risk factor *GBA1* gene (n=10),^15,29^ and in the monogenic *LRRK2* (n=1)^30^ or *PRKN* genes (n=1)^28^ (Supplementary Table 3). Among the patients with *GBA1* variants, only two were able to have their LEDD substantially reduced (by ≥30% or ≥50%) post-operatively. Although the sample size of *GBA1*-PD patients was relatively small, median % LEDD reductions were significantly less in these patients vs. those without *GBA1* variants at both T2 (by 19.0% [31.5%] vs. 52.9% [38.9%], P=0.002) and T3 (20.0% [50.5%] vs. 48.3% [39.6%], P=0.015), compared to T1 (with no significant between-group difference in baseline LEDD). *GBA1*-variant carriers accounted for two of the patients with dementia (severe in one [PD-1414], with significant cognitive problems starting within a few months post-operatively in her 50s (13 years after PD diagnosis) despite undergoing otherwise uncomplicated DBS (first-pass placements of the cranial electrodes and no peri-operative brain haemorrhage);^27^ and moderately severe in the other [PD-0203], occurring within four years post-operatively in his 50s [17 years post-diagnosis]);^15^ and one of the patients (PD-2045) having problematic gambling and attempting suicide (described above). A good DBS outcome was obtained in the patient with PARK-*PRKN*,^28^ whereas the benefit was small-to-medium only in the patient with *LRRK2* p.R1441C.^30^

The “Asian” *LRRK2* risk variants p.R1628P and p.G2385R were detected in 10.3% (15/145) and 7.6% (10/132) of patients, respectively. The median LEDD at baseline and the median % LEDD reductions at T2 and T3 (vs. T1) among these patients did not differ significantly vs. those without the *LRRK2* risk variants (data not shown).

### 3.4 Dystonia subgroup

There were five patients, with: tardive dystonia (n=2); familial pure generalized dystonia (n=1); familial generalized dystonia-parkinsonism (n=1); and idiopathic Meige syndrome (n=1). All underwent bilateral GPi DBS. Outcomes were mixed: the patient with the generalized tardive dystonia had a medium-to-large benefit; the patient with familial pure generalized dystonia a medium benefit; the patient with idiopathic Meige syndrome a small-to-medium benefit, and the remaining two patients (with familial generalized dystonia-parkinsonism and tardive segmental dystonia) experiencing small benefits only.

## 4. DISCUSSION

DBS can often dramatically improve the motor function and quality of life of patients suffering from movement disorders, including PD and dystonias, that are not responding satisfactorily to medical therapy.^1,2^ However, largely due to its relatively high cost (especially the cost of the device),^11^ the vast majority of patients worldwide are unable to benefit from a technological “advance” that has been deployed in clinical practice for more than 20 years.^12^ In our study, less than one-fifth of DBS cases were government-funded, reflecting the fact that in developing countries, healthcare costs are often largely borne by patients and families. ^11,21^

Another factor that limits the more widespread application of DBS is the fact that the outcome is highly dependent on the skills of the DBS team, particularly in patient selection, the accuracy of neurosurgical targeting, and post-operative management. These take time to develop, as does the trust and confidence among the community of patients and clinicians in the safety and efficacy of the procedure done in the local setting.^11^

Over time, and in parallel with the advent of rechargeable IPGs providing an improved benefit-to-cost ratio, our centre has been able to provide DBS treatment to a growing number of patients, with overall good outcomes as evidenced by complication and clinical efficacy metrics that are on par with those reported internationally. For example, rates of symptomatic intracranial haemorrhage were 1.9% (vs. 0.6-6.0%); SSI 4.3% (vs. 0-15.2%); and wire fracture 0.6% (vs. 0.7-4.4%).^31–33^ In PD patients (by far the commonest indication for DBS in our series), LEDD reduction was ≥50% and ≥30% in 42.2% and 69.8%, respectively, at 6-12 months post-DBS.

Despite the safety and efficacy of DBS, however, outcomes can sometimes be less than satisfactory, both for patients and their families, as well as for the treating clinicians. This may arise because of unrealistic expectations;^34^ surgical complications; ongoing issues with levodopa-related complications (e.g., dyskinesias that fail to abate despite LEDD reduction^6^); or cognitive-behavioural, balance or other life-limiting problems caused by lesioning from the brain surgery, off-target stimulation effects, disease progression, and/or comorbidities.

Thus, one therapeutic strategy advocated in recent years has been to offer DBS at earlier disease stages in younger patients with: (i) lower complication risks; (ii) still-intact functional (e.g., occupational) capacities; and (iii) more prolonged benefit (before this becomes overshadowed by disease progression or comorbidities).^4,5^ This approach was evident in our cohort, where there has been a trend of decreasing disease duration at the time of DBS surgery, corresponding to an increasing number of cases with short-duration PD.

The validity of this strategy seems to be supported by our safety and efficacy data – besides having acceptable complication rates, our short-duration patients had on average greater and more sustained LEDD reductions compared to patients with standard-duration PD. Importantly, while misdiagnosis of early-stage atypical parkinsonism for PD is commonly cited as a reason to avoid early DBS,^9,26^ none of our short-duration patients had their diagnosis reassigned during follow-up. We acknowledge that these disorders may sometimes not declare themselves until much later, e.g., in a series from the Mayo Clinic, several patients undergoing DBS for “PD” received a revised diagnosis of multiple system atrophy as late as 15, 16 or 17 years after disease onset.^35^

Currently, the literature regarding the relationship between PD genetic variants and presentation for, and response to, DBS is still limited. Our rate of monogenic and *GBA1*-related PD in the subset of patients tested (12/61=19.7%) was comparable to the findings of DBS cohorts in the United States^36^ (26.5% of 100 EOPD patients) and the United Kingdom^37^ (28.7% of 94 unselected patients) (to our knowledge, similar studies have not yet been performed in Asian populations). The frequencies of *GBA1* variants (16.4%, vs. 12.1%^36^ and 17.0%^37^) were also comparable. Studies indicate that *GBA1* variants are associated with less favourable outcomes after DBS,^16,38^ including worse cognitive decline, axial motor features, function, and quality of life, and less LEDD reduction, consistent with our preliminary observations. Indeed, the 19.0-20.0% LEDD reduction in our patients was remarkably similar to the meta-analysed figure of 22%, involving 30 *GBA1* variant carriers from white populations.^38^ The “severe” p.L483P variant in particular has been associated with worse disease progression in white populations,^39^ and the findings in this DBS cohort (where p.L483P accounted for 90.0% of the pathogenic/likely pathogenic *GBA1* variants, vs. 18.8-25.0% in previous reports^36,37^) suggest that this may also be the case in Asians.^12^

The rates of *LRRK2* Asian risk variants in our cohort (p.R1628P, 10.3%; p.G2385, 7.6%) were comparable to overall (non-DBS) Asian PD populations (each variant being present in ≍5-10% of patients^12^). We are aware of only one study, involving a Han Chinese PD cohort, that systematically studied p.G2385R frequency in patients undergoing DBS and found this to be relatively high (8/57=14.0%),^40^ suggesting that in some populations its presence could be associated with motor complications^41^ requiring DBS. The presence of Asian *LRRK2* variants did not seem to adversely affect treatment response in our patients, similar to the findings of Chen et al.^40^ Higher-powered, systematic studies will allow more definitive conclusions to be drawn regarding the roles of *LRRK2* and other gene variants (as well as, potentially, polygenic risk scores) in the context of DBS, particularly in patients lacking pathogenic or high-risk PD variants.^18,19,42^

The sample size for dystonia in this study was very small, allowing only tentative comments to be made. DBS efficacy for tardive syndromes remains to be demonstrated in larger randomized studies;^43^ in line with this, our two patients with tardive dystonia had mixed outcomes and continued to have significant disability and require botulinum toxin injections post-operatively. Certain monogenic dystonias (e.g., DYT-*TOR1A* and DYT-*KMT2B*) respond favourably to DBS,^44^ however, in our movement disorder clinics, we have detected DYT-*KMT2B* in only two patients^45^ and DYT-*TOR1A* in none, potentially explaining the limited benefit of DBS in our dystonia cases.

Interestingly, although DBS is an established treatment for essential tremor (ET), no patient in our cohort underwent (or was referred for) DBS for ET. ET is commonly diagnosed, but it seems that this is rarely sufficiently severe in Malaysian patients to be considered for neurosurgical treatment, again suggesting potential ethno-geographic differences in the expression of movement disorders.^29,46^ In a survey of experts from Asia, it was reported that only a “very low” number of essential tremor patients undergo functional neurosurgery in India, although in Japan 210 cases were treated with focused ultrasound lesioning over a three-year period (2016-2019).^11^

Our study has several strengths and limitations. It was single-centre, and the sample size of 155 patients is relatively small compared to reports from developed nations. On the flip side, patients were managed in a consistent manner by a small dedicated team of clinicians, which we believe increased the reliability of between-group comparisons (e.g., short-vs. standard-duration PD, or *GBA1* variant-positive vs. variant-negative subgroups). The study also represents the largest published on DBS from Southeast Asia and provides a valuable picture of DBS practice in an under-represented region of the world. Another limitation was the lack of systematic, quantitative documentation of a range of disease-related variables (motor and non-motor function, functional abilities, quality of life, satisfaction with treatment, etc.) before and after DBS. The gold standard to objectively verify motor improvement after DBS is to evaluate patients after overnight withdrawal of PD medications, however, we lacked the human resources to routinely do this. Furthermore, since the evidence base for DBS efficacy is considered well established,^24^ these assessments were deemed to pose unnecessary physical and financial hardship to patients.^12,21^ On the positive side, the study provides data that are reflective of real-world clinical practice, and included all operated patients, not just those who were willing or able to participate in more demanding research protocols. Although conveying only a partial picture of a patient’s overall condition post-DBS, dopaminergic medication reduction is a good indication of STN DBS efficacy in improving PD symptoms, and attenuates medication-related adverse effects including dyskinesias and hyperdopaminergic behaviours.^2,18,47^ Indeed, the rates of overt medication-related neuropsychiatric complications in our patients (with generally more advanced disease) were low.

In conclusion, we described real-world experience of DBS in a developing country and showed that the procedures were safe and effective. A notable aspect of our cohort was the inclusion of a substantial number of patients with a short duration of PD, in whom good clinical outcomes were seen, with larger and more sustained reductions in PD medication requirement, and none subsequently developing atypical parkinsonism. Our findings also provide Asian-relevant genotype-phenotype insights, particularly with regards to *GBA1* and *LRRK2* variants that are relatively commonly encountered. Last but not least, the study highlights inequity of access to a potentially life-changing treatment, calling for urgent consideration and action from the clinical-scientific and patient-support communities and partners in industry.

## DISCLOSURES

### Ethical Compliance Statement

We confirm that we have read the Journal’s position on issues involved in ethical publication and affirm that this work is consistent with those guidelines.

### Financial Disclosures for the Previous 12 months

SYL receives grants from the Michael J. Fox Foundation and the Ministry of Education Malaysia Fundamental Research Grant Scheme. He serves on the Editorial Board for Neurotorium. He has received honoraria for talks sponsored by Medtronic, and the International Parkinson & Movement Disorder Society (MDS). AHT receives grants from the Michael J. Fox Foundation, Ministry of Education Malaysia Fundamental Research Grant Scheme, and Toray Science Foundation Science & Technology Grant. She has received honoraria for talks sponsored by Sanofi, and the International Parkinson & Movement Disorder Society (MDS). KAM is a proctor for Medtronic and has received honoraria from Medtronic.

## Supporting information

Mal-DBS Suppplemental Materials

## Data Availability

All data produced in the present study are available upon reasonable request to the authors.

## Acknowledgements

The authors gratefully acknowledge funding from The Ministry of Higher Education Malaysia Fundamental Research Grant Scheme (FRGS/1/2020/SKK0/UM/01/2) and University of Malaya Parkinson’s Disease and Movement Disorders Research Program (PV035-2017). This project was supported by the Global Parkinson’s Genetics Program (GP2). GP2 is funded by the Aligning Science Across Parkinson’s (ASAP) initiative and implemented by The Michael J. Fox Foundation for Parkinson’s Research (https://gp2.org). For a complete list of GP2 members see https://gp2.org. The authors also gratefully acknowledge Medtronic staff Mr. Brian Kah Chun Hor, Mr. Lybron Lee Kian Pooi, and Ms. Amira Aina Hasbullah for providing DBS-related technical support to patients and their families.

## AUTHOR ROLES

1. Research project: A. Conception, B. Organization, C. Execution
2. Statistical Analysis: A. Design, B. Execution, C. Review and Critique
3. Manuscript Preparation: A. Writing of the first draft, B. Review and Critique.

AMD: 1B-C, 2A-C, 3A-B

SYL: 1A-C, 2C, 3A-B

KAM: 1B-C, 3B

AHT: 1B-C, 2A, 3B

All other authors: 1C, 3B

