## Supplementary material for "Real-World Experience of Deep Brain Stimulation Surgery in a Developing Southeast Asian Country": Mal-DBS Suppplemental Materials

**Supplementary Table 1. Procedure-related complications after deep brain stimulation and their management and outcomes**

| Complication | Number of patients | Management & Outcome |
|---|---|---|
| Intracerebral hemorrhage | 2 | Medical management<br><br>In the 1st patient (Parkinson's disease [PD] diagnosed at 60s; underwent surgery around 5 years later), there was some functional recovery from the stroke (e.g., became ambulant again), and the IPG was subsequently implanted; unfortunately, this was complicated by SSI leading to explantation of the cranial leads and implantable pulse generator (IPG)<br><br>In the 2nd patient (PD diagnosed at 50s; underwent surgery around 9 years later), the IPG was never implanted due to poor functional status (non-recovery from the stroke) |
| Subdural hemorrhage (SDH) | 1 | In this patient (PD diagnosed at 60s; underwent surgery around 3 years later), the left STN lead implantation was complicated by SDH and she underwent immediate surgical evacuation of the SDH upon detection intra-operatively (the patient had complained of headache, without neurologic deficits); the right STN lead and IPG were successfully implanted 1 month later |
| Surgical site infection (SSI) | 7 (including the 1st patient above with intracerebral hemorrhage) | Antibiotics and surgical debridement (all 7); the cranial leads (12 leads) and/or IPG (5 IPGs) had to be explanted in 6 patients |
| Skin erosion | 3 | Antibiotics, surgical debridement and explantation of IPG in 2 patients and intracranial leads in 1 patient, with successful re-implantation subsequently (all 3) |
| Lead fracture (resulting from major fall down the stairs) | 1 | The patient presented with muscle contractions likely due to capsular spread of current, and worsening parkinsonian features on the left side of the body<br>Right STN DBS lead successfully re-implanted |
| Seizure | 2 (including the 2nd patient above with intracerebral hemorrhage) | Medical management with control of seizures |

**Supplementary Table 2. Clinico-demographic features of patients with short-duration Parkinson's disease, and indications for deep brain stimulation.**

| Cohort Patient Designation | Age Range at Diagnosis (years) | Interval between diagnosis and surgery (years) | Disease Duration at Surgery (years) | Indication(s) |
|---|---|---|---|---|
| 1. PD-1343 | 40-49 | 4.1 | 4.1 | Troublesome motor fluctuations |
| 2. PD-2143 | 50-59 | 4.0 | 4.0 | Troublesome motor fluctuations |
| 3. PD-0876 | 50-59 | 4.5 | 4.5 | Troublesome motor fluctuations |
| 4. PD-1107 | 40-49 | 3.0 | 3.0 | Troublesome motor fluctuations and dyskinesias |
| 5. PD-1610 | 50-59 | 2.5 | 2.5 | Troublesome motor fluctuations |
| 6. PD-1916 | 60-69 | 4.4 | 4.4 | Troublesome motor fluctuations and dyskinesias |
| 7. PD-1550 | 50-59 | 4.1 | 4.1 | Troublesome tremors |
| 8. PD-1860 | 40-49 | 3.6 | 3.6 | Troublesome motor fluctuations |
| 9. PD-1926 | 60-69 | 2.6 | 2.6 | Troublesome motor fluctuations and tremors |
| 10. PD-1747 | 30-39 | 2.8 | 2.8 | Troublesome motor fluctuations and intolerance to PD medications |
| 11. PD-1734 | 40-49 | 2.7 | 2.7 | Troublesome motor fluctuations |
| 12. PD-1618 | 60-69 | 4.1 | 4.1 | Troublesome motor fluctuations and dyskinesias |
| 13. PD-2172 | 50-59 | 2.8 | 2.8 | Troublesome motor fluctuations |
| 14. PD-2157 | 60-69 | 2.3 | 2.3 | Troublesome motor fluctuations |
| 15. PD-2241 | 50-59 | 4.7 | 4.7 | Troublesome motor fluctuations |
| 16. PD-1859 | 40-49 | 3.3 | 3.3 | Troublesome motor fluctuations and dyskinesias |
| 17. PD-1790 | 60-69 | 3.4 | 3.4 | Troublesome tremor and intolerance to PD medications |
| 18. PD-2424 | 60-69 | 3.6 | 3.6 | Troublesome motor fluctuations |
| 19. PD-1364 | 30-39 | 4.7 | 4.7 | Troublesome motor fluctuations and dyskinesias |
| 20. PD-2617 | 40-49 | 2.3 | 2.3 | Troublesome dyskinesias |
| 21. PD-2113 | 30-39 | 2.8 | 2.8 | Troublesome motor fluctuations |
| 22. DBS-132 | 60-69 | 3.0 | 3.0 | Troublesome motor fluctuations |
| 23. PD-2371 | 40-49 | 4.8 | 4.8 | Troublesome motor fluctuations and intolerance to PD medications (psychosis) |
| 24. PD-2534 | 50-59 | 3.9 | 3.9 | Troublesome dyskinesias |
| 25. PD-3074 | 40-49 | 2.7 | 2.7 | Troublesome motor fluctuations |

**Supplementary Table 3. Post-operative reduction of levodopa-equivalent daily dose (LEDD) in Parkinson's disease (PD) patients harbouring pathogenic/likely pathogenic PD gene variants and/or "Asian" *LRRK2* risk variants.** For easier visualization, cases shaded in: grey harboured pathogenic or likely pathogenic *GBA1* variants; green those with ≥30% post-operative reduction in LEDD (with some cases exceeding 50%); red those where 30% reduction was not achieved.

| Patient | Ancestry | Genetic variant | T2 vs. T1 | | T3 vs. T1 | |
|---|---|---|---|---|---|---|
| | | | LEDD reduction ≥30% | LEDD reduction ≥50% | LEDD reduction ≥30% | LEDD reduction ≥50% |
| 1. PD-0135 | Chinese | *GBA1* p.L483P (P) | Yes | No | No | No |
| 2. PD-1484 | Chinese | | No | No | No | No |
| 3. PD-1598 | Chinese | | No | No | No | No |
| 4. PD-1859 | Chinese | | No | No | No | No |
| 5. PD-2045 | Chinese | | No | No | Yes | No |
| 6. PD-2887 | Chinese | | Pending | Pending | Pending | Pending |
| 7. PD-2188 | Chinese | *GBA1* p.L483P (P) + *LRRK2* p.G2385R | No | No | No | No |
| 8. PD-1414[27] | Chinese | *GBA1* RecNcil (P) + *LRRK2* p.R1628P | N/A | N/A | No | No |
| 9. PD-1428 | Chinese | | Yes | No | Yes | Yes |
| 10. PD-0203[15] | Indian | *GBA1* p.N227S (LP) + *PRKN* exon 5 deletion | No | No | No | No |
| 11. PD-0219[28] | Chinese | *LRRK2* p.R1441C (P) | Yes | No | No | No |
| 12. PD-1010[26] | Chinese | *PRKN* compound heterozygous exon 1 deletion (P) + p.Glu309* (LP) *in trans* + *LRRK2* p.R1628P | Yes | Yes | Pending | Pending |
| 13. PD-0114 | Chinese | *LRRK2* p.R1628P | No | No | No | No |
| 14. PD-0155 | Chinese | | Yes | No | Yes | Yes |
| 15. PD-1367 | Mixed[#] | | Yes | Yes | Yes | Yes |
| 16. PD-1548 | Chinese | | Yes | Yes | No | No |
| 17. PD-1650 | Chinese | | Yes | Yes | Yes | Yes |
| 18. PD-1795 | Chinese | | Pending | Pending | Pending | Pending |
| 19. PD-2292 | Chinese | | Yes | No | No | No |
| 20. PD-2371 | Chinese | | Yes | Yes | Pending | Pending |
| 21. PD-2379 | Chinese | | Yes | Yes | Yes | No |
| 22. PD-2534 | Chinese | | Pending | Pending | Pending | Pending |
| 23. PD-2859 | Chinese | | Yes | Yes | No | No |
| 24. PD-0198 | Chinese | | N/A | N/A | Yes | Yes |

| | | | | | | |
|---|---|---|---|---|---|---|
| 25. PD-1091 | Chinese | *LRRK2* p.G2385R | Yes | Yes | No | No |
| 26. PD-1475 | Chinese | | No | No | No | No |
| 27. PD-1597 | Chinese | | Yes | Yes | Yes | Yes |
| 28. PD-1839 | Chinese | | No | No | Yes | No |
| 29. PD-1890 | Chinese | | No | No | Yes | No |
| 30. PD-1918 | Chinese | | No | No | Yes | Yes |
| 31. PD-2188 | Chinese | | No | No | No | No |
| 32. PD-2597 | Chinese | | Pending | Pending | Pending | Pending |
| 33. PD-1610 | Chinese | *LRRK2* p.R1628P + *LRRK2* p.G2385R | Yes | No | No | No |

*GBA1* variants were found in 9/57 (15.8%) of the early-onset Parkinson's disease (PD) patient subgroup who underwent genetic testing, and in 1/10 (10%) of the short-duration PD patient subgroup. References for four patients with monogenic PD who were reported previously are indicated by superscripts in the first column. RecNcil is a recombinant allele represented by the co-inheritance of p.L483P(;)A495P(;)V499V. [#]Mixed=Chinese + Indigenous/Bidayuh. LP=Likely pathogenic; N/A=Not available; P=Pathogenic; T1=Pre-DBS; T2=Within 6-12 months post-DBS; T3=At last hospital visit.

**APPENDIX A.**

Studies in Southeast Asia using (Deep Brain Stimulation[MeSH Major Topic]) AND (country[Affiliation])

1. Singapore (3 studies, 93 patients)
    1. Karthick PA, Wan KR, An Qi AS, Dauwels J, King NKK. Automated detection of subthalamic nucleus in deep brain stimulation surgery for Parkinson's disease using microelectrode recordings and wavelet packet features. J Neurosci Methods. 2020 Sep 1;343:108826. doi: 10.1016/j.jneumeth.2020.108826. Epub 2020 Jul 2. PMID: 32622981. (n=26)
    2. Ng JH, See AAQ, Xu Z, King NKK. Longitudinal medication profile and cost savings in Parkinson's disease patients after bilateral subthalamic nucleus deep brain stimulation. J Neurol. 2020 Aug;267(8):2443-2454. doi: 10.1007/s00415-020-09741-3. Epub 2020 May 4. PMID: 32367298. (n=56)
    3. Ang J, Zhang JJY, Yam M, Maszczyk T, Ng WH, Wan KR. Clinical Application of a Stereotactic Frame-Specific 3D-Printed Attachment for Deep Brain Stimulation Surgery. World Neurosurg. 2023 Feb;170:e777-e783. doi: 10.1016/j.wneu.2022.11.121. Epub 2022 Nov 29. PMID: 36455844. (n=11)
2. Philippines (3 studies, 41 patients)
    1. Abejero JEE, Jamora RDG, Vesagas TS, Teleg RA, Rosales RL, Anlacan JP, Velasquez MS, Aguilar JA. Long-term outcomes of pallidal deep brain stimulation in X-linked dystonia parkinsonism (XDP): Up to 84 months follow-up and review of literature. Parkinsonism Relat Disord. 2019 Mar;60:81-86. doi: 10.1016/j.parkreldis.2018.09.022. Epub 2018 Sep 21. PMID: 30262378. (n=11)
    2. Diestro JDB, Vesagas TS, Teleg RA, Aguilar JA, Anlacan JP, Jamora RDG. Deep Brain Stimulation for Parkinson Disease in the Philippines: Outcomes of the Philippine Movement Disorder Surgery Center. World Neurosurg. 2018 Jul;115:e650-e658. doi: 10.1016/j.wneu.2018.04.125. Epub 2018 Apr 28. PMID: 29709756. (n=17)
    3. Dannug AT, Gabriel FGC, Macias MCYL, Diesta CCE. Impact of deep brain stimulation on quality of life and motor symptoms in Parkinson's disease and X-linked dystonia parkinsonism: The Philippine experience. Parkinsonism Relat Disord. 2021 Jun;87:92-97. doi: 10.1016/j.parkreldis.2021.04.026. Epub 2021 May 6. PMID: 34015695. (n=13)
    4. [Brüggemann N, Domingo A, Rasche D, Moll CKE, Rosales RL, Jamora RDG, Hanssen H, Münchau A, Graf J, Weissbach A, Tadic V, Diesta CC, Volkmann J, Kühn A, Münte TF, Tronnier V, Klein C. Association of Pallidal Neurostimulation and Outcome Predictors With X-linked Dystonia Parkinsonism. JAMA Neurol. 2019 Feb 1;76(2):211-216. doi: 10.1001/jamaneurol.2018.3777. PMID: 30508028; PMCID: PMC6439955.] - done in Luebeck
3. Thailand (3 studies, 84 patients)
    1. Nunta-Aree S, Sitthinamsuwan B, Boonyapisit K, Pisarnpong A. SW2-year outcomes of subthalamic deep brain stimulation for idiopathic Parkinson's disease. J Med Assoc Thai. 2010 May;93(5):529-40. PMID: 20524438. (n=27)

2. Phokaewvarangkul O, Boonpang K, Bhidayasiri R. Subthalamic deep brain stimulation aggravates speech problems in Parkinson's disease: Objective and subjective analysis of the influence of stimulation frequency and electrode contact location. Parkinsonism Relat Disord. 2019 Sep;66:110-116. doi: 10.1016/j.parkreldis.2019.07.020. Epub 2019 Jul 16. PMID: 31327627. (n=50)
3. Nunta-aree S, Nimmannitya P, Sitthinamsuwan B, Pisarnpong A, Boonyapisit K. Clinical Outcome of Pallidal Deep Brain Stimulation for Various Types of Dystonia. J Med Assoc Thai 2017;100:103. (n=7)

4. Malaysia (1 patient)

1. Idris Z, Ghani AR, Mar W, Bhaskar S, Wan Hassan WN, Tharakan J, Abdullah JM, Omar J, Abass S, Hussin S, Abdullah WZ. Intracerebral haematomas after deep brain stimulation surgery in a patient with Tourette syndrome and low factor XIIIA activity. J Clin Neurosci. 2010 Oct;17(10):1343-4. doi: 10.1016/j.jocn.2010.01.054. PMID: 20620064. (n=1)

**APPENDIX B - Deep Brain Stimulation Surgery Protocol at the University of Malaya**

**Patient selection.** Patients are initially assessed and medically managed by movement disorders neurologists (SYL and AHT). Patients deemed potentially suitable for DBS are referred to the DBS neurosurgeon (KAM). The typical indication for DBS in patients with Parkinson's disease (PD) is the presence of troublesome motor response complications (motor fluctuations and/or dyskinesias). Cases of troublesome tremor that are not satisfactorily controlled by medication treatment, or where PD medications are associated with intolerable adverse effects, are also considered. Patients with dementia are excluded. Neuropsychiatric problems such as depression, psychosis or impulsive-compulsive behaviours (ICBs) are not excluded, provided the condition is not florid and considered to pose an imminent risk to the patient during the post-operative period (e.g., triggering suicide or self-injurious behaviours). There is no minimal duration of PD for patients to be considered eligible, as long as there is no suspicion of an atypical parkinsonian disorder.

Cases of dystonia, tremor, and other movement disorders (e.g., tics/Tourette's syndrome) are also considered for DBS if symptoms are troublesome and have not responded satisfactorily to medication treatment.

**Pre-operative work-up.** Patients are typically admitted to hospital for two days, to undergo a non-contrasted brain MRI scan, neuropsychiatric assessment, anaesthestic evaluation, and medical consultation as needed (e.g., for cardiological issues). For patients with PD, a levodopa challenge is performed to document a substantial improvement in the medication-ON vs. medication-OFF state. Details of the DBS procedure, including the anticipated benefits and potential risks, are further discussed with the patient and family/significant other(s).

**DBS operation.** Patients are admitted on the day prior to the DBS surgery and those with PD have their dopaminergic medications taken off for ≥12 hours prior to the surgery (in some cases where, for example, minimal parkinsonian rigidity was detected previously during the medication-OFF state, patients may be advised to taper and come off longer-acting dopamine agonist medications over the preceding 1-2 weeks). Antithrombotics are withheld when feasible for one week, and traditional medications/supplements for at least one month.

On the day of the procedure, the patient undergoes a non-contrasted cranial CT scan with the CRW® stereotactic frame. Images are fused with the pre-operative MR images and coordinates are obtained in accordance to radiologic planning on the structural target done previously using Stereoplan® software in the Brain Lab®/Medtronic® planning station.

For PD patients, the scalp incisions and twist drill craniotomy are done under local anesthesia. Light sedation is achieved with Target Controlled Infusions (TCI, Minto Model) of remifentanil. In very anxious patients, dexmedetomidine (Precedex®) infusion is sometimes used after discussion with the neurosurgeon and neurologist. Labetalol infusion is used to manage intra-operative hypertension.

Microelectrode recording is utilized to localise and map the subthalamic nucleus. This is followed by macrostimulation using the microelectrode cannula and then macrostimulation with the DBS lead (usually up to 4.5V, at 60microseconds pulse width and 130Hz frequency). Intraoperative clinical assessments are done by the movement disorders neurologist, assessing for response to stimulation (typically improvement in upper limb rigidity, ± tremor where this is a prominent feature), and for the occurrence of stimulation-related adverse effects (with a focus on dysconjugate eye deviation, capsular pulling and speech disturbance). In the vast majority of cases, the cranial leads are implanted "first pass" to minimize the possibility of complications. Antibiotic is administered prophylactically (intravenous cefuroxime 1.5g followed by 750mg at the time of

IPG implantation). The implantable pulse generator (IPG) is then internalized under general anaesthesia.

For dystonia patients, DBS lead implantation and IPG internalization are all done under general anesthesia.

A post-operative brain CT is done immediately after the surgery to assess lead placement and to rule out intracranial bleeding.

Patients are typically discharged from hospital the following day, once the post-operative antibiotic course (intravenous cefuroxime 750mg 3 doses post-operatively) is completed, with neurosurgical clinic follow-up 7-14 days later for wound monitoring and removal of sutures.

**Post-operative management.** DBS programming for PD patients is typically first carried out 4-6 weeks after surgery, once any lesional effects have subsided and patients are back to their pre-operative clinical status and have resumed their usual PD medications. Patients come to clinic in the medication-OFF state (stopped for ≥12 hours). Each contact is tested in turn for efficacy and side effects, and the contact with the largest therapeutic window (i.e., lowest threshold for benefit and highest threshold for adverse effects) and/or eliciting the largest clinical improvement is then activated in a single monopolar configuration, at 1.0 or 1.5V, 60microseconds and 130Hz. Subsequent programmings are undertaken on a case-by-case basis, according to the patient's response, logistic factors, and ability to self-program. In recent years, with the availability of a patient-controlled programmer, many adjustments (typically gradual increases in voltage, by 0.1 or 0.2V each time) are now made at home by the patient/family, guided by the neurologist via email, instead of the patient having to come in for frequent adjustments (e.g., at 1-8-weekly intervals).

Patients are advised to gradually reduce their PD medications as their PD symptoms improve in conjunction with the gradual increases in the stimulation parameters. However, complete cessation of PD medication is usually discouraged, to avoid problems such as depression or apathy. In general, COMT inhibitors are taken off first, followed by levodopa reduction (especially if dyskinesias are a problem), although sometimes dopamine agonist therapy is prioritized for reduction/cessation (e.g., due to the presence of ICBs or psychosis, or where their relatively higher cost is an issue).
